# Molecular architecture of human atherosclerosis revealed through integrative human genetics

**DOI:** 10.1101/2025.03.06.25323506

**Authors:** Bassim El-Sabawi, Michael Betti, Phillip Lin, Xiaoning Huang, Namju Kim, Mohammad Yaser Anwar, Andrew S. Perry, B. Lakshitha A. Perera, Priya Gajjar, Laura A. Colangelo, Kaushik Amancherla, Quanhu Sheng, Shilin Zhao, Lindsay Stolze, Eric Farber-Eger, Joshua M. Landman, Patricia E. Miller, Gabrielle Y. Liu, Suman Das, Quinn S. Wells, James G. Terry, Donald Lloyd-Jones, Saumya Das, Sadiya S. Khan, Kari E. North, Jennifer Below, Matthew Nayor, Ravi Kalhan, John Jeffery Carr, Eric R. Gamazon, Ravi V. Shah

## Abstract

Current genetic discovery methods are largely restricted to profiling circulating molecules or genetic architecture, limited in use of tissue-based molecular genetics to identify pathogenic and therapeutic targets. Here, we leverage a multi-level genetic discovery platform integrating population-level proteomics with functional genomic analyses based on human coronary artery tissue to reveal determinants of coronary disease susceptibility. Using aptamer-based proteomics (≈7,000 aptamers) across ≈3,000 individuals, we first identified the circulating proteome of prevalent and incident coronary artery calcium (CAC)—a sensitive marker of subclinical coronary artery disease (CAD)—with causal implication in calcified plaque formation or disease phenotypes via parallel genetic approaches (Mendelian randomization, MR) and proteome-wide association (PWAS). Identified proteins specified pathways of extracellular matrix remodeling, immune cell function, lipid metabolism, and inflammation. To resolve findings at a coronary tissue level, we performed the largest to date coronary artery-specific transcriptome-wide association study for CAC (TWAS; based on RNA-seq from 268 human coronary arteries) in >35,000 individuals, demonstrating enrichment of targets from the circulating proteome with supportive evidence by traditional MR approaches (*NOTCH3, SPINK2, S100A12, RPP25, OAF, HS6ST3, TNFSF12, GPC6*), several implicated in CVD-adjacent biological mechanisms. Phenome-wide association and single cell transcriptomics in human coronary arteries across atherosclerosis implicated targets in tissue-specific disease mechanisms. Finally, using coronary artery-specific functional genomic annotations of chromatin structure, conformation, and accessibility, we identified trans regulation of two of these genes (*GPC6*, *RPP25*) by CAC GWAS-significant SNPs, resolving targets for previously “orphan” genome-wide significant loci. These multi-level findings furnish a resource for pathobiology of CAC, atherosclerosis, and establish an adaptable framework applicable to all organ systems to parse precision targets for prevention, surveillance, and therapy of cardiovascular disease.

## INTRODUCTION

Atherosclerotic cardiovascular disease (CVD) is the most common cause of cardiovascular demise worldwide^1^. CVD has origins decades before the occurrence of key clinical events, including sudden cardiac death and myocardial infarction^2^. Despite ongoing, broad pharmacologic and behavioral efforts to mitigate risk, progressive worsening in dysmetabolic phenotypes has promoted an increase in early CVD, with nearly 50% of CVD mortality under age 65 in some reports^3,4^. Accordingly, defining underlying targetable CVD mechanisms at an early subclinical stage is a major goal in modern precision cardiovascular prevention. Coronary artery calcification (CAC)—a touchstone phenotype of early CVD susceptibility with therapeutic implication^5^—represents a culmination of interconnected molecular events within endothelium and the vascular wall involving local and systemic inflammation and immune activation and metabolic-oxidative stress^6^. Molecular studies of early coronary disease progression in humans have focused on relating accessible genetic or circulating metabolic states (via metabolites, proteins, peripheral blood transcriptomes)^7–9^ to CAC or clinical CVD. Nevertheless, examining underlying determinants of coronary artery disease in a *tissue-specific fashion*—heretofore limited in large populations due to inaccessibility of coronary tissue^6^—is critical to identify and interrupt early, intervenable changes in human CVD. Furthermore, despite innovation in human genetic resources in the last 10 years spanning tissue transcription and epigenetics, the application of these technologies in a multi-dimensional fashion to prioritize targets of <u>functional</u> interest in CVD remains limited.

Here, we leverage a translational genetic discovery platform integrating proteomics, genomics, coronary artery (bulk tissue and single cell) transcription, and coronary artery epigenomics to identify causal determinants of human coronary artery disease. We first identified a human proteomic signature of CAC through population-level proteome-wide association with prevalent and 10-year incident coronary artery calcium phenotypes in ≈3,000 individuals. To parse potential causal drivers of coronary phenotypes from proteomics, we deployed two complementary methods—a protein quantitative trait loci (pQTL) based Mendelian randomization (MR) and a proteome-wide association study (PWAS) approach—to map targets from circulation with substantial genetic evidence to CVD. In parallel, we performed the largest transcriptome-wide association study (TWAS) of CAC in >35,000 individuals using genetic models of gene expression developed in human coronary arteries, identifying population-level circulating proteomic associations with convergent evidence from genetically determined expression in the causal tissue. We next leveraged an epigenomic atlas of coronary artery regulatory elements and chromatin conformation capture (Hi-C) data to identify genes consistently associated with CAC across the multiple approaches (coronary TWAS, MR, proteomics) as the distal (*trans*) regulatory targets of GWAS-implicated variants. Targets identified from this multi-parametric approach were finally tested for differential expression across human coronaries with and without disease via single-cell transcriptomics. This approach represents a comprehensive, multi-dimensional paradigm to human coronary atherosclerosis, incorporating both circulating and tissue-derived molecular architecture of disease liability, prioritizing causal, tissue-relevant determinants of coronary disease.

## RESULTS

### Characteristics of proteomic discovery cohorts

Our overall study scheme is shown in **Figure 1**. Clinical characteristics of the study populations (Coronary Artery Risk Development in Young Adults; CARDIA; N=2,971; Framingham Heart Study; FHS; N=573) are in **Table 1**. CARDIA participants at Year 25 had a mean age of 50.2±3.6 years, with approximately even distribution by sex and self-identified race (56% women, 46% Black) and prevalent cardiometabolic risk (mean systolic blood pressure 119±15 mmHg, 8% diabetes, mean BMI 30.3±7.1 kg/m^2^) and CAC (≈29%). A subgroup of participants with baseline proteomics and without prevalent CAC at Year 25 had computed tomography performed ≈10 years later (Year 35 study visit in CARDIA; N=793), with an incident CAC rate of ≈31%. FHS participants were similar in age and sex, with a low prevalence of clinical cardiovascular disease (≈4%) at the time of proteomics, with a high rate of incident CAC (assessed ≈20 years after proteomics, ≈69%).

**Figure 1.**
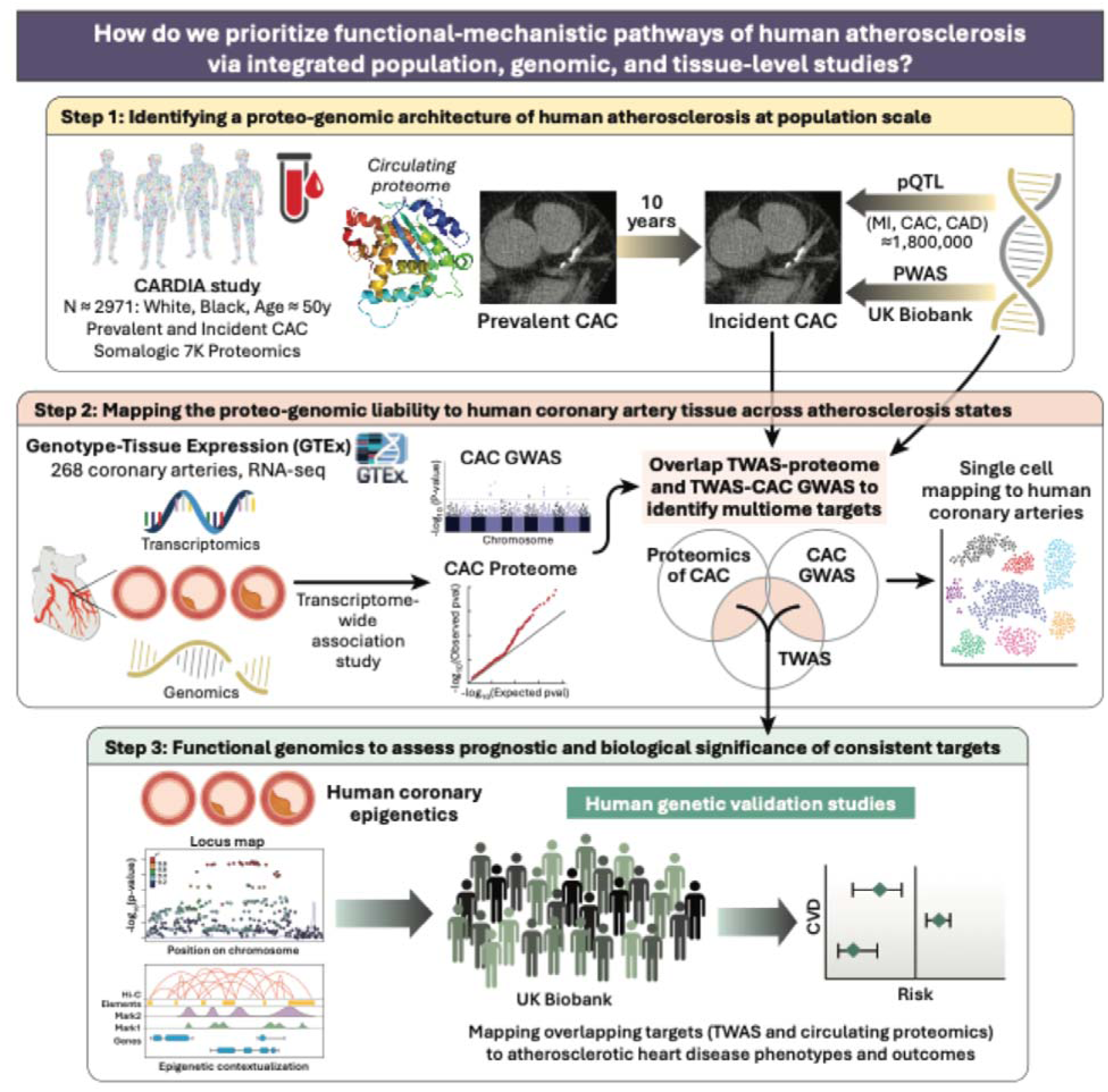
Study scheme illustrating multi-level approach for discovery in human coronary disease.

**Table 1.** Characteristics of study samples. Panel **A** shows data from CARDIA study from the Year 25 examination. Panel **B** shows data from FHS.

| Covariate | Derive (N=2,087) | Validate (N=884) |
| --- | --- | --- |
|  | Mean (SD) / N (%) | Mean (SD) / N (%) |
| Age | 50.1 (3.6) | 50.3 (3.6) |
| Female | 1,166 (55.9%) | 495 (56.0%) |
| Black | 961 (46.0%) | 419 (47.4%) |
| Body mass index (kg/m<sup>2</sup>) | 30.3 (7.1) | 30.1 (7.2) |
| Systolic blood pressure (mmHg) | 118.8 (15.4) | 118.2 (15.2) |
| Smoking status |  |  |
| Current smoker | 570 (27.5%) | 221 (25.1%) |
| Quit within a year | 111 (5.4%) | 60 (6.8%) |
| Never or quit over a year | 1,392 (67.1%) | 598 (68.0%) |
| Diabetes (fasting glucose ≥ 126 mg/dl or on medications) | 166 (8.0%) | 74 (8.4%) |
| Total cholesterol (mg/dL) | 192.4 (36.9) | 191.3 (36.6) |
| Total high-density lipoprotein cholesterol (mg/dL) | 57.5 (17.8) | 58.3 (17.9) |
| Any CVD disease by Y25 | 65 (3.1%) | 32 (3.6%) |
| Coronary artery calcium (AU) | 48.0 (210.9) | 48.6 (265.6) |
| Coronary artery calcium >0 (AU) | 634 (30.4%) | 222 (25.1%) |
| Abdominal aorta calcium (AU) | 219.7 (666.1) | 185.4 (581.0) |
| Abdominal aorta calcium > 0 (AU) | 1,082 (52.9%) | 457 (53.5%) |

**B**
| Covariate | N=573<br>Mean (SD) / N (%) |
| --- | --- |
| Age | 51 (9) |
| Female | 339 (59.2%) |
| Non-White | 2 (0.3%) |
| Body mass index (kg/m<sup>2</sup>) | 27 (5) |
| Current smoker | 67 (11.7%) |
| Systolic blood pressure (mmHg) | 122 (17) |
| Treated for hypertension | 47 (8.2%) |
| Diabetes | 17 (3.0%) |
| Total cholesterol (mg/dL) | 202 (35) |
| Total high-density lipoprotein cholesterol (mg/dL) | 52 (15) |
| Treated for lipids | 26 (4.5%) |
| Any CVD by exam 5 | 9 (1.6%) |
| Any CVD by CT scan date* | 55 (9.6%) |
| CAC > 0 (AU) | 395 (69%) |
| CAC (mean +/- SD) (AU) | 324 (705) |
| AAC > 0 (AU) | 493 (87%) |
| AAC (mean+/- SD) (AU) | 2,463 (3,535) |

\*CT scan was performed a median of 17.4 years after proteomics were measured

### Identifying and characterizing the circulating proteomic architecture of coronary artery calcification

We identified 136 aptamers (representing 131 unique proteins) associated with presence <u>and</u> extent of CAC across discovery and validation sets within CARDIA (**Figure 2A**; presence of CAC, 186 aptamers; extent of CAC, 201 aptamers; regression summaries in **Supplementary Table 1**). Proteins with highest effect sizes in regression exhibited strong biological plausibility across known mechanisms of vascular remodeling (top 20 aptamers for CAC extent in validation set shown in **Table 2**), including fibrosis and inflammatory mechanisms (GDF-15^10^, CDCP1^11^, GSN^12^, TSP-2^13^, chemokines), oxidative lipid metabolism (CILP2^14^), extracellular matrix remodeling and signaling (MMP-7, MMP-12^15^, TIMP-1, integrins^16^), calcification (Notch 1^17^, ARHGAP36^18^), and general metabolism (GIP^19^). Importantly, a majority of identified protein associations (though biologically plausible) had not been previously widely reported in human coronary disease or calcification, including mechanisms of protein catabolism (UBE2G2^20^), signal transduction (RHG36), macrophage efferocytosis and lipid metabolism (TREM2^21^), endothelial cell states (EGFR^22^), and extracellular matrix metabolism (PCOC1).

**Figure 2.**
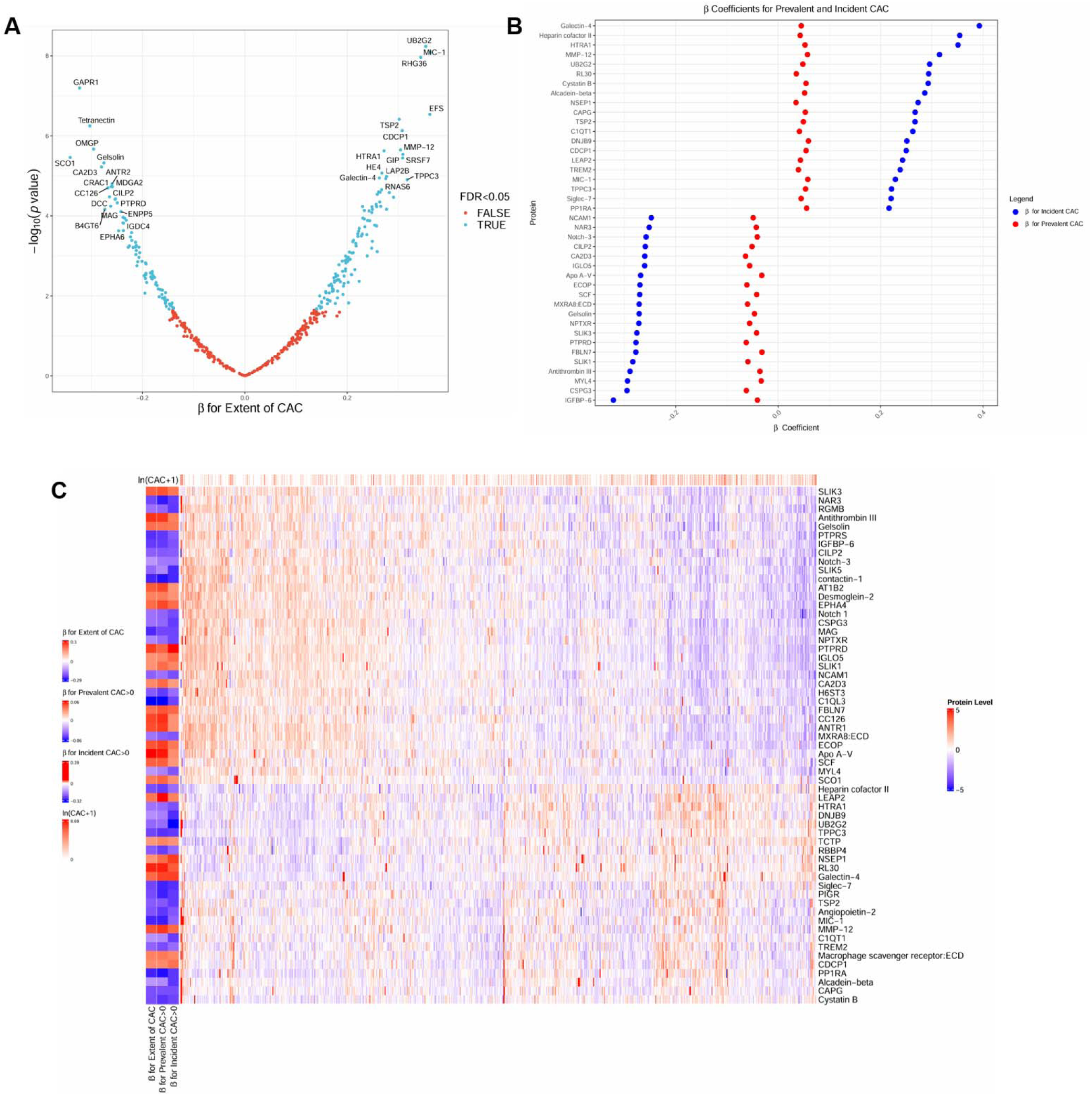
Circulating proteomic architecture of human coronary calcification. (A) Volcano plot displaying the relationships of aptamers with the extent of CAC in the validation set. (B) Plot showing beta coefficients for prevalent CAC>0 in the derivation sample and incident CAC>0 for the aptamers with the top 20 and bottom 20 beta coefficients for incident CAC>0. (C) Heatmap illustrating the log_2_-transformed, standardized, and winsorized levels of aptamers across all participants. The heatmap features 59 aptamers associated with the presence and extent of CAC at Year 25 in CARDIA, which were also associated with incident CAC at Year 35. The heatbars on the left represent the beta coefficients from models assessing prevalent CAC>0 at Year 25, extent of CAC at Year 25, and incident CAC at Year 35. The heatbar on the top shows ln(CAC+1) at Year 25 per subject.

**Table 2.** Existing molecular evidence of proteins with the top 20 absolute effect size for extent of CAC in the validation sample that are associated with both presence and extent of CAC.

| Gene/Protein | CAC Effect | Molecular Evidence |
| --- | --- | --- |
| **Inflammation and fibrosis** |  |  |
| GDF15 (Growth/differentiation factor 15) | ↑ | A member of the TGF-β cytokine family; serves as a robust and independent biomarker for predicting disease progression and mortality in individuals with coronary artery disease.<sup>101</sup> |
| EFS (Embryonal Fyn-associated substrate) | ↑ | CRK-associated substrate adaptor protein; modulates T-cell function via negative selection; implicated in immune function and autoimmunity<sup>102</sup> |
| GSN (Gelsolin) | ↓ | Related to inflammation, lipid metabolism, and angiogenesis<sup>12</sup>; Expression is reduced arteriosclerosis compared to normal tissue |
| CDCP1 (CUB domain-containing protein 1) | ↑ | Regulates cardiac fibroblasts proliferation, mitosis, and implicated in myocardial fibrosis<sup>11</sup> |
| CTSD (Cathepsin D) | ↑ | An enzyme found in macrophage lysosomes that plays a role in modifying low-density lipoproteins, facilitating their uptake by macrophages and contributing to foam cell formation<sup>103</sup> |
| UBE2G2 (Ubiquitin-conjugating enzyme E2 G2) | ↑ | Involved in protein degradation; differentially expressed in early coronary atherosclerosis<sup>104</sup> and knockdown suppressed atherosclerotic plaque formation in mice<sup>20</sup> |
| GLIPR2 (Golgi-associated plant pathogenesis-related protein 1) | ↓ | Negative regulator of autophagy; upregulated in fibrosis<sup>105</sup> |
| **Calcification** |  |  |
| ARHGAP36 (Rho GTPase-activating protein 36) | ↑ | Involved in regulating bone development and metabolism; overexpression linked to heterotopic ossification<sup>18</sup> |
| **Extracellular Remodeling** |  |  |
| ADAMTSL2 (ADAMTS-like protein 2) | ↑ | Regulates extracellular matrix deposition and remodeling; negative regulator of TGF-β and attenuates myofibroblast differentiation and mitigates fibrosis in cardiomyopathies<sup>106</sup> |
| CLEC3B (Tetranectin) | ↓ | Regulator of the fibrinolysis and proteolytic system; decreased circulating levels in patients with coronary artery disease<sup>107</sup> |
| MMP12 (Macrophage metalloelastase) | ↑ | Involved in extracellular remodeling; overexpressed in atherosclerotic plaques and increased plasma levels associated with atherosclerotic burden and prevalent cardiovascular disease<sup>108,109</sup> |
| THBS2 (Thrombospondin-2) | ↑ | Modulates cell-matrix interactions; uncertain role in atherosclerosis development, TSP-2 mRNA is not expressed in the endothelial cells that line atheroma<sup>110</sup> |
| **Metabolism** |  |  |
| GIP (gastric inhibitory polypeptide) | ↑ | Regulates vasodilation, vascular leukocyte adhesion, inflammation, and lipid metabolism; exerts both anti- and pro-atherosclerotic effects depending on the environment<sup>111</sup> |
| **Other** |  |  |
| SCO1 (Protein SCO1 homolog, mitochondrial) | ↓ | Mitochondrial protein that regulates cellular copper importation; involved in oxidative stress response<sup>112</sup> |
| OMG (Oligodendrocyte-myelin glycoprotein) | ↓ | Cell adhesion molecule required for myelination in the central nervous system<sup>113</sup>. |
| CACNA2D3 (Voltage-dependent calcium channel subunit alpha-2/delta-3) | ↓ | A component of the voltage-gated calcium channel complex |
| TMPO (Lamina-associated polypeptide 2, isoforms beta/gamma) | ↑ | Plays a role in maintaining nuclear architecture and facilitating post-mitotic nuclear reassembly |
| CSTB (Cystatin B) | ↑ | Inhibitor of cysteine proteases |
| TRAPPC3 (Trafficking protein particle complex subunit 3) | ↑ | Plays a role in transporting proteins within the Golgi and endoplasmic reticulum<sup>114</sup> |
| SRSF7 (serine/arginine-rich splicing factor 7) | ↑ | Regulate precursor RNA splice site recognition and spliceosome assembly; implicated in malignant cell proliferation<sup>115</sup> |

To address epidemiologic limitations on cross-sectional associations (reverse causation), we quantified the relation of our prevalent CAC proteome with incident development of CAC ≈10 years later in a subsample of CARDIA without CAC at time of proteomics (N = 793; **Figure 2B-C, Supplementary Table 2**). Among aptamers related to prevalence and extent of CAC, we observed strong consistency in directionality of effect (129/136 aptamers) with incident CAC, including 59 aptamers that were associated with incident CAC at 10 years. In addition, we examined proteomic associations with CAC measured ≈20 years after proteomics in FHS, observing overall consistency in effect directionality and size (for extent of CAC: Pearson r = 0.47, p<0.001), with 649 of 1,035 aptamers exhibiting concordant directionality of effect (**Supplementary Figure 1**; full regression results in **Supplementary Table 3**).

### Human genetic prioritization of mediators of cardiovascular disease in the human CAC proteome

To increase confidence in the targetability of select mediators and their causal relevance to human CVD, we next assessed a potential causal role for proteins implicated by CAC through two complementary approaches: (1) Mendelian randomization (MR) and (2) proteome-wide association studies (PWAS). For MR, we used established protein quantitative trait loci (pQTL) for proteins associated with presence <u>and</u> extent of CAC in CARDIA (100 included that satisfied quality control thresholds; see **Methods**). We observed broad consistency between MR-based genetic estimates of effect on three major disease conditions (CAC, coronary artery disease, and myocardial infarction; spanning ≈1.8 million individuals, study characteristics shown in **Supplementary Table 4**) and proteomic-phenotype associations from CARDIA (**Figure 3A**, full results in **Supplementary Table 5**), several of which have not been widely reported in coronary vascular calcification in humans. Proteins with both proteomic and genetic evidence in favor of increased calcification and clinical risk included mediators involved in central mechanisms of metabolism and inflammation, including *GABARAPL1*^23^ (autophagy), *S100A9/*calgranulin B (inflammatory signaling/atherosclerosis in murine models^24^), *VEGFA*^25,26^ (angiogenesis and lipid handling), *C1QTNF1*^27^ (macrophage activation). Importantly, several genes with evidence of protection against endpoints at a proteomic and genomic level had not been broadly implicated in calcification in humans, including proteins implicated in central nervous system development and physiology (*SLITRK1*^28,29^: noradrenergic system development; *IGLON5*^30^; *NTRK3*), extracellular matrix metabolism (*HS6STG3*, *ADAM23*) and fibroblast development (*FGFR1*).

**Figure 3.**
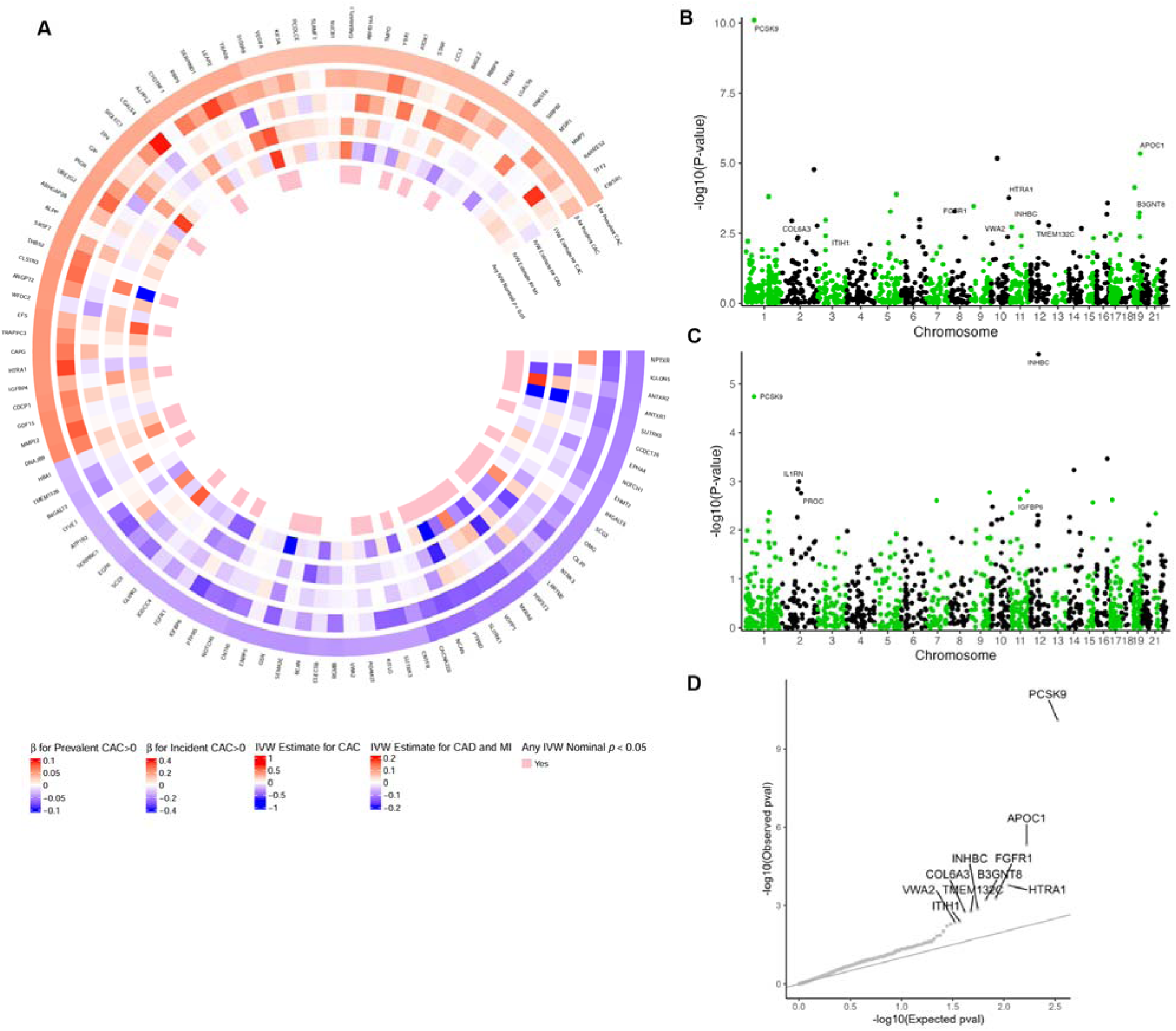
Proteo-genomics of human coronary calcification. (A) Circular heatmap depicting 100 proteins associated with both the presence and extent of CAC and with available protein quantitative trait loci (pQTLs) as genetic instruments for Mendelian randomization analysis of CAC, coronary artery disease (CAD), and myocardial infarction (MI) from genome-wide association studies (GWAS). The heatmap illustrates the relationships between beta coefficients for proteomic associations with prevalent CAC>0 and incident CAC>0, as well as inverse variance weighted (IVW) effect sizes for CAC, CAD, and MI, demonstrating overall concordance. The inner most tracks highlight genes with IVW nominal p-values < 0.05 with any outcome (CAC, CAD, or MI). (B) PWAS for coronary atherosclerosis in UK Biobank (UK Biobank code: I9_CORATHER). (C) PWAS for myocardial infarction in UK Biobank (UK Biobank code: I9_MI). For (B) and (C), labeled proteins were associated with presence and extent of CAC in CARDIA (derivation P<0.05) and the PWAS (P<5×10^-3^). Of 753 proteins with derivation P<0.05 for the proteomic association in CARDIA, 316 had available genetic models of protein abundance and could be tested in PWAS. (D) Quantile-quantile plot showing enrichment for (PWAS) associations of genetically determined protein abundance with coronary atherosclerosis in the UK Biobank (UK Biobank code: I9_CORATHER) among the proteins in circulation associated with presence and extent of CAC in CARDIA (derivation P<0.05).

In a parallel approach, using genetic data from 7,213 individuals with proteomics and genomic data, we developed a pipeline for PWAS of circulating protein abundance across 4,657 aptamers (Somalogic; see **Methods**). We estimated the association of genetically determined protein abundance with atherosclerotic disease phenotypes in a cohort of 361,194 UK Biobank participants. At a 5% Bonferroni threshold, we observed four proteins with significant evidence for involvement of genetically determined protein abundance in atherosclerosis (*PCSK9*, *APOC1, HTRA1, and FGFR1*; **Figure 3B**) and two for myocardial infarction (*PCSK9*, *INHBC*; **Figure 3C**; full results in **Supplemental Table 6**) among those proteins that had suitable PWAS models (i.e., P<0.05, cross-validation *r*=0.10, 1,340 unique proteins) and evidence for association with CAC in CARDIA proteomics. Notably, the directly measured, CAC-associated proteins were substantially enriched for genetically determined CAC associations (**Figure 3D**). Circulating PCSK9 expression is linked to greater severity and vulnerability of coronary plaque^31^, with successful PCSK9 interruption linked to lower vascular event rates^32^. Both *CXCL12* (an inflammatory chemokine) and *APOC1* (lipoprotein metabolism) have been implicated in human GWAS or functional studies of atherosclerosis^33,34^. *INHBC* (implicated in adipocyte metabolism) appears to link cardiometabolic health to coronary risk, with higher levels related to higher coronary risk^35^.

### Coronary artery-specific transcriptome-wide association studies (TWAS) and single cell transcription implicates both known and novel targets in human atherosclerosis

Circulating proteome-wide association—either via genetic instruments (PWAS) or directly measured proteins (CARDIA)—cannot resolve target tissue-specific causal mechanisms of disease pathogenesis. TWAS in a mechanistically relevant tissue may address this limitation, estimating genetically determined transcript expression within a trait-related tissue (in this case, human coronary artery) to use in conjunction with cognate phenotype GWAS data (in this case, invivo human measured calcified coronary plaque or CAC). TWAS in such mechanistically relevant tissues enhances specificity of identified GWAS associations. Furthermore, GWAS variant associations are frequently non-coding and can be within genomically “dense” loci or gene “deserts,” limiting precise inference on which SNP-linked gene is functional.

Therefore, we constructed genetic models for gene expression in 268 human coronary arteries (see **Methods**), mapping these models to the largest extant GWAS of CAC^36^ to identify gene-level associations with CAC. Coronary artery expression of genes associated with CAC in TWAS (**Figure 4A**; full results in **Supplemental Table 7**) included known (*PHACTR1*^37^, *MRAS*^37^*, MORF4L1-ADAMTS7*^38^) and not previously widely implicated targets (*GIGYF1*, *DMPK*, *RPL9*). *PHACTR1* (phosphatase and actin regulator-1) and *MRAS* demonstrated strong association with CAC, consistent with pathogenic roles in endothelial dysfunction, CAC, or inflammatory cell function^37,39,40^. Interestingly, *MORF4L1* and *ADAMTS7* are adjacent at 15q25.1 with the lead SNP in an intronic region of *MORF4L1*^38^, leading to difficulty in resolving the actual causal disease gene. While only *ADAMTS7* has wide support in models of atherosclerosis^38^, our TWAS results (based on genetically determined RNA expression in coronary artery) along with MR to account for LD contamination implicated both genes in CAC pathogenesis. Other genes identified in coronary TWAS of CAC displayed phenotypic association with anatomically or functionally adjacent processes, though not directly with atherosclerosis itself (*GIGYF1*: type 2 diabetes^41^; *DMPK*: cardiac conduction^42^; *RPL9*: ribosomal metabolism^43^). Importantly, despite differences in origin, we observed, among the genes encoding proteins significantly associated with CAC in CARDIA, an enrichment for coronary artery TWAS associations with CAC (**Figure 4B**), including both known mediators of vascular dysfunction or plaque homeostasis (*NOTCH3*^44^, *TNFSF12*^45^, *S100A12*^46^) as well as several not previously widely related to vascular disease.

**Figure 4.**
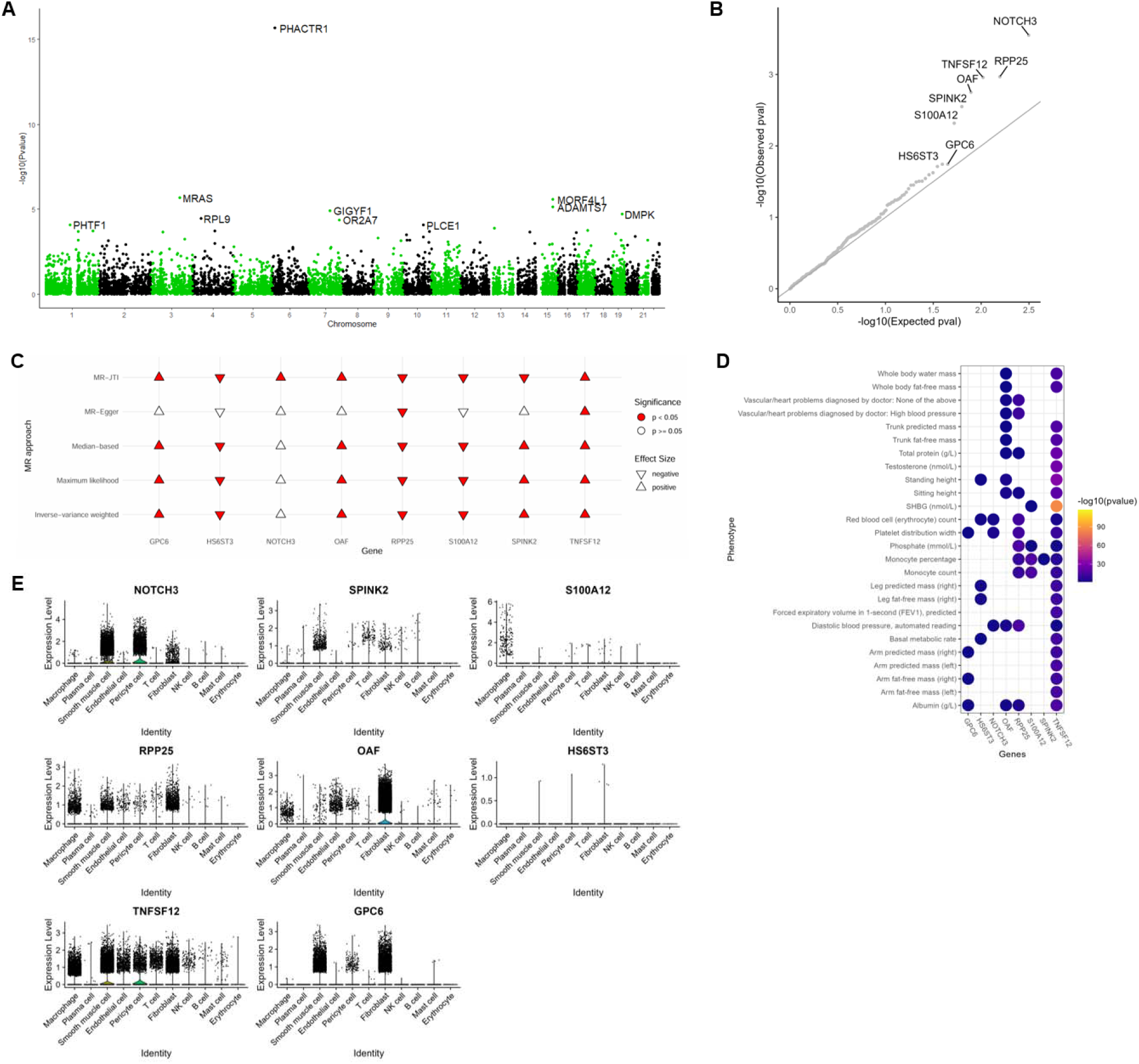
Human coronary transcriptome-wide association study of CAC and single cell transcription. (A) Transcriptome-wide association study of CAC (N=35,776) using genetic models of gene expression in human coronary samples. (B) Quantile-quantile plot showing enrichment for TWAS associations among the proteins associated with presence and extent of CAC in CARDIA (derivation P<0.05). (C) Causal inference of CAC TWAS associations using Mendelian randomization. Causal inference was run using five different Mendelian randomization (MR) approaches: MR-JTI, MR-Egger, median-based, maximum likelihood, and inverse-variance weighted. Direction of effect is indicated by the direction of each triangle (right side up for positive and upside down for negative), and MR analyses that were statistically significant for causality are indicated in red. (D) Heatmap showing associations of the genetically determined expression of the 8 genes across heritable phenome captured in the UK Biobank. All TWAS associations have P-value < 0.05. (E) Gene expression profile from single-cell transcriptomics. Using single-cell RNA sequencing data in human atherosclerosis, we identified the expression profile of the 8 genes across the cell types.

Overall, our approach prioritized 8 genes with multi-level support across proteomic association with CAC and human coronary artery TWAS of CAC. In comprehensive causal analyses of CAC using 5 different MR approaches (each with its own set of strong assumptions; **Methods**), all 8 genes (*NOTCH3*, *SPINK2*, *S100A12*, *RPP25*, *OAF*, *HS6ST3*, *TNFSF12, GPC6*) had supportive evidence for causal associations (**Figure 4C**). For 5 of these genes (*SPINK2*, *RPP25*, *OAF*, *HS6ST3*, *GPC6*), the direction of effect observed in MR analyses was consistent with the direction of the circulating proteomic association with CAC. Of note, none of the 8 genes have been therapeutically targeted in CAD, and less than half have been implicated in CVD directly (as noted above for *NOTCH3*, *TNFSF12*, *S100A12*)^44–46^. Genetic variants in *HS6ST3*—a gene whose product has been implicated in protein-extracellular matrix interactions—have been linked to CAC in some settings^47^, though implications in human atherosclerosis have not been investigated. In addition, while not previously directly implicated in CVD, both *GPC6* and *RPP25* have support: *GPC6* blocks canonical Wnt signaling^48^, a pathway central to vascular calcification^49,50^, and *RPP25* (a part of ribonuclease P) has been implicated in genetic studies of cardiometabolic disease^51^ and in scleroderma (a disease marked by multi-organ calcification and fibrosis). In phenome-wide association studies (PheWAS) across the heritable phenome (heritability h^2^ P < 0.05) in UK Biobank using genetically determined expression in coronary artery, many of the 8 prioritized genes implicated vascular, inflammatory, and obesity-related phenotypes (**Figure 4D**). The 8 genes were expressed across broad cell types in human coronary arteries^52^ implicated in mechanisms of progressive calcification, including vascular inflammation, fibrosis, and smooth muscle cell remodeling (**Figure 4E**). Collectively, these results demonstrate genetic support for proteomic targets from circulation and implicate a target gene expression within human coronary arteries in CAC pathogenesis.

### Coronary artery-specific functional genomics identifies genome-wide significant *trans* regulatory elements for targets with multi-level evidence in human CAC

We then conducted extensive characterization of the nature of the genetic regulation of these 8 genes to gain crucial insights into disease mechanisms, with critical methodological relevance for the functional interpretation of GWAS findings, using a comprehensive array of epigenomic, chromatin accessibility and transcription factor binding assays in human coronary artery. We identified a total of 11,795 SNPs interrogated in the CAC GWAS that overlap a coronary artery regulatory element in contact (Hi-C) with one of the 8 genes. Two loci (rs9515203 and rs28610385) attained genome-wide significance (P<5×10^-8^; **Figure 5A**). The SNP rs9515203 lies in an intron of *COL4A2* and falls in an insulator—characterized by enrichment of CTCF and cohesin complex subunits RAD21 and SMC3—in 3D contact with the distal target *GPC6*, a gene over 10 MB upstream on chromosome 13 (**Figure 5B-C**). Similarly, the SNP rs28610385 lies in an intron of *ADAMTS7* and falls in a transcribed gene body element—characterized by accessible chromatin (as measured by ATAC-seq) as well as enrichment of histone marks H3K36me3, H3K79me2, and H4K20me1—in 3D contact with the distal target gene *RPP25* (**Figure 5D-E**).

**Figure 5.**
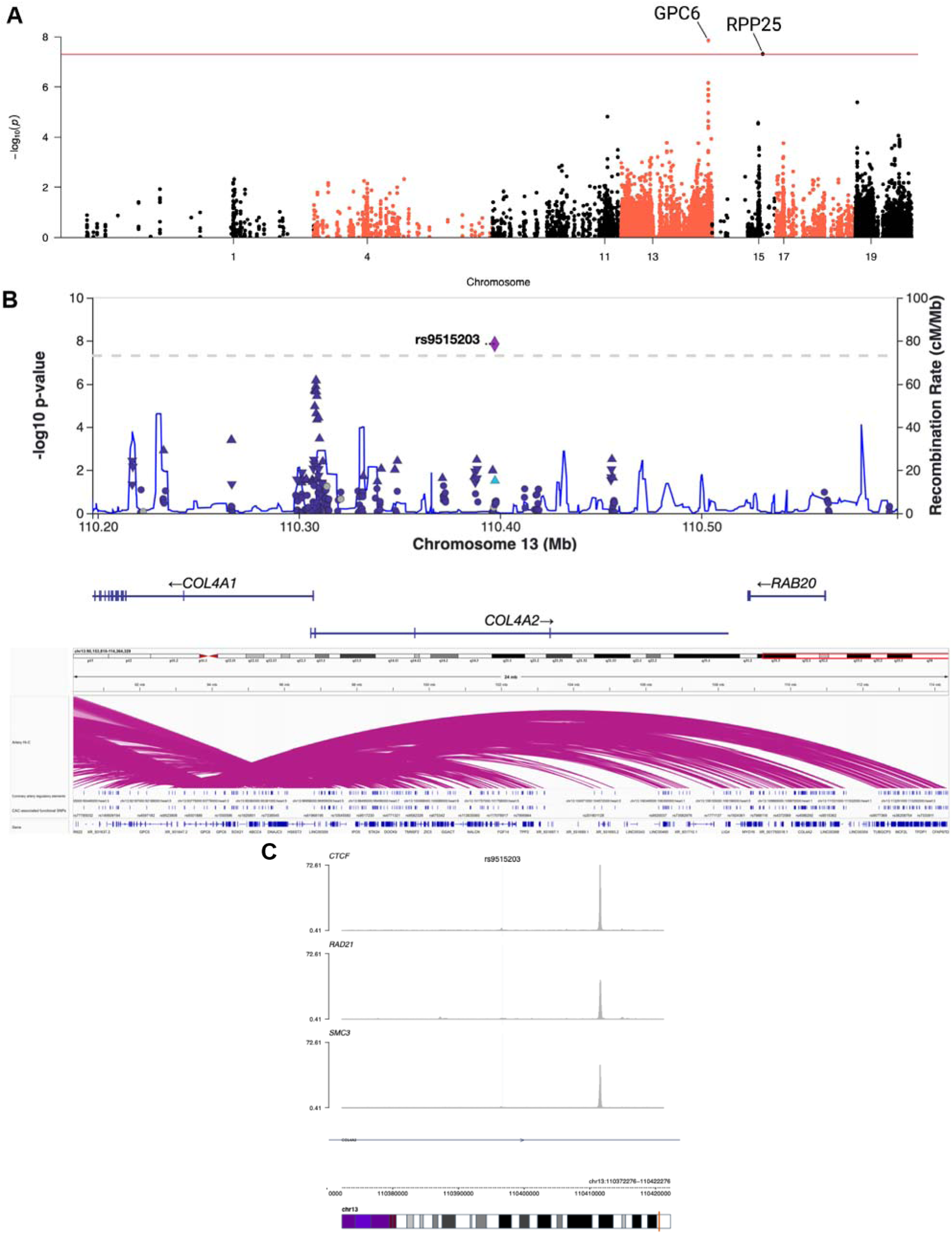
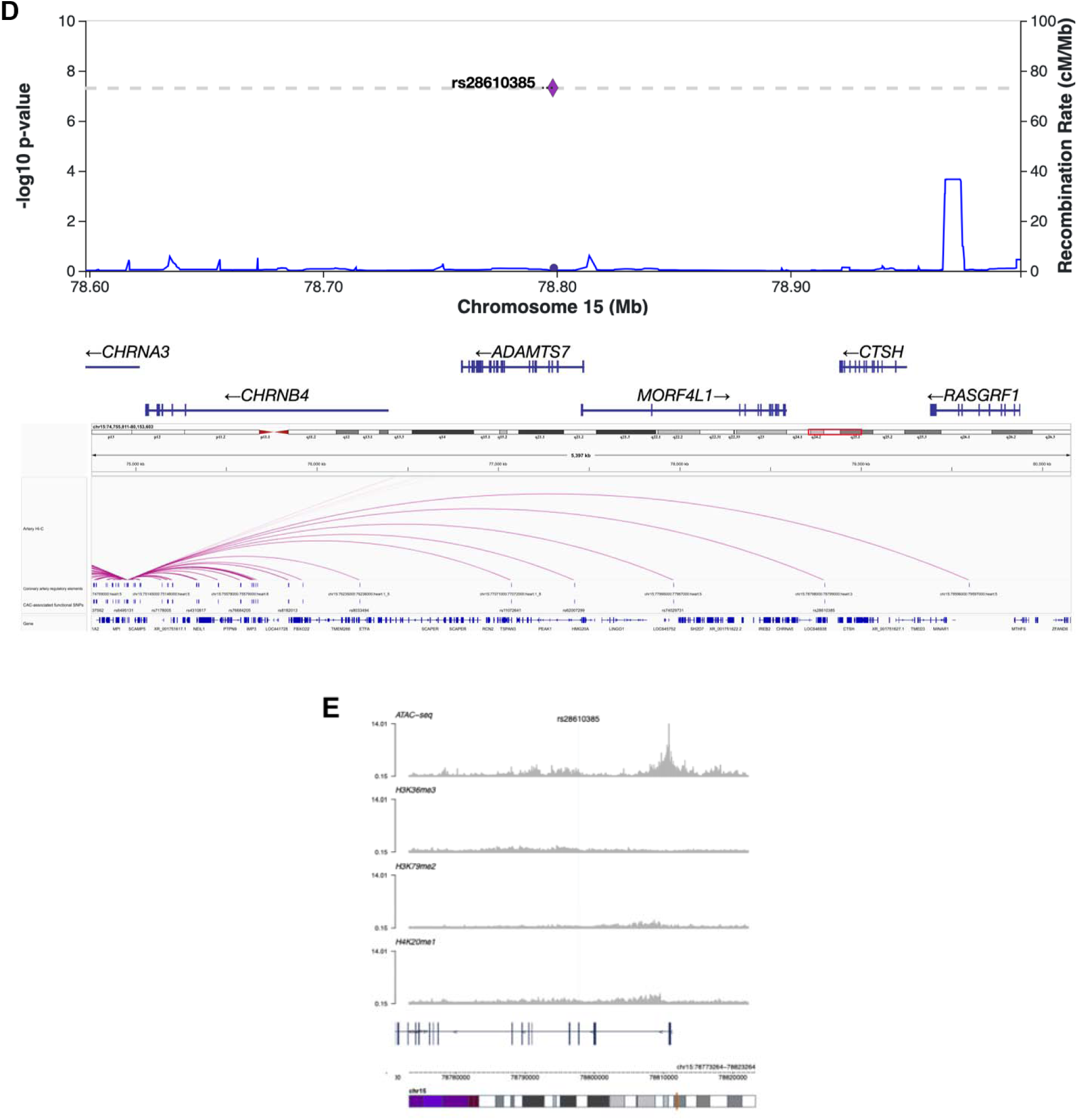
Human coronary functional genomics studies. (A) CAC GWAS SNPs linked to one of eight associated genes from convergent proteome and TWAS associations. We identified a total of 11,795 CAC GWAS SNPs that overlapped a coronary artery regulatory element in 3D contact (Hi-C) with one of the eight CAC-associated genes (*HS6ST3*, *GPC6*, *S100A12*, *SPINK2*, *OAF*, *TNFSF12*, *RPP25*, and *NOTCH3*). Of these SNPs, two (rs9515203 linked to *GPC6* and rs28610385 linked to *RPP25*) were genome-wide significant (*p* < 5 x 10^-8^). (B) **LocusZoom plot of rs9515203. *Top:*** Depicted are all functional SNPs (i.e., overlapping regulatory elements in coronary artery) linked to CAC-associated genes in coronary artery that neighbor rs9515203. ***Bottom:* Zoomed out plot of rs9515203 contact locus.** SNP rs9515203 overlaps with an insulator in coronary artery (designated chr13:110397000:110398000). This element falls within an intron of *COL4A2* and makes distal 3D contact (Hi-C) with *GPC6*, a gene over 10 Mb upstream on chromosome 13. (C) Chromatin signature around rs9515203 implicates an insulator in coronary artery. Regulatory elements in this class are characterized by enrichment of CTCF and cohesin complex subunits RAD21 and SMC3. (D) ***Top:* LocusZoom plot of rs28610385.** Depicted are all functional SNPs (i.e., overlapping regulatory elements in coronary artery) linked to CAC-associated genes in coronary artery that neighbor rs28610385. ***Bottom:* Zoomed in plot of rs28610385 contact locus.** SNP rs28610385 overlaps with a transcribed gene body element in coronary artery (designated chr15:78798000:78799000). This element partially overlaps an intron of *ADAMTS7* and makes distal 3D contact (Hi-C) with *RPP25*. (E) Chromatin signature around rs28610385 shows a transcribed gene body element in coronary artery. Regulatory elements in this class are characterized by chromatin accessibility (in this case, measured by ATAC-seq), as well as the histone marks H3K36me3, H3K79me2, and H4K20me1. We observed enrichment of these 3 chromatin marks throughout the locus of rs28610385 in the coronary artery epigenome.

Using the single-cell transcriptomics data for additional support of the gene regulatory relationships for the GWAS-implicated CAC-associated variants, *GPC6* (glypican-6) was significantly differentially expressed in *atherosclerosis* in fibroblasts (P=4.87×10^-25^), pericytes (P=7.63×10^-7^), and smooth muscle cells (P=6.29×10^-5^). *RPP25* was differentially expressed *in atherosclerosis* in fibroblasts (P=0.004) and endothelial cells (P=0.01; **Figure 6**). Collectively, these results highlight the role of *trans* (distal) genetic regulation, by GWAS-implicated variants, of tissue-specific gene expression in disease pathogenesis.

**Figure 6.**
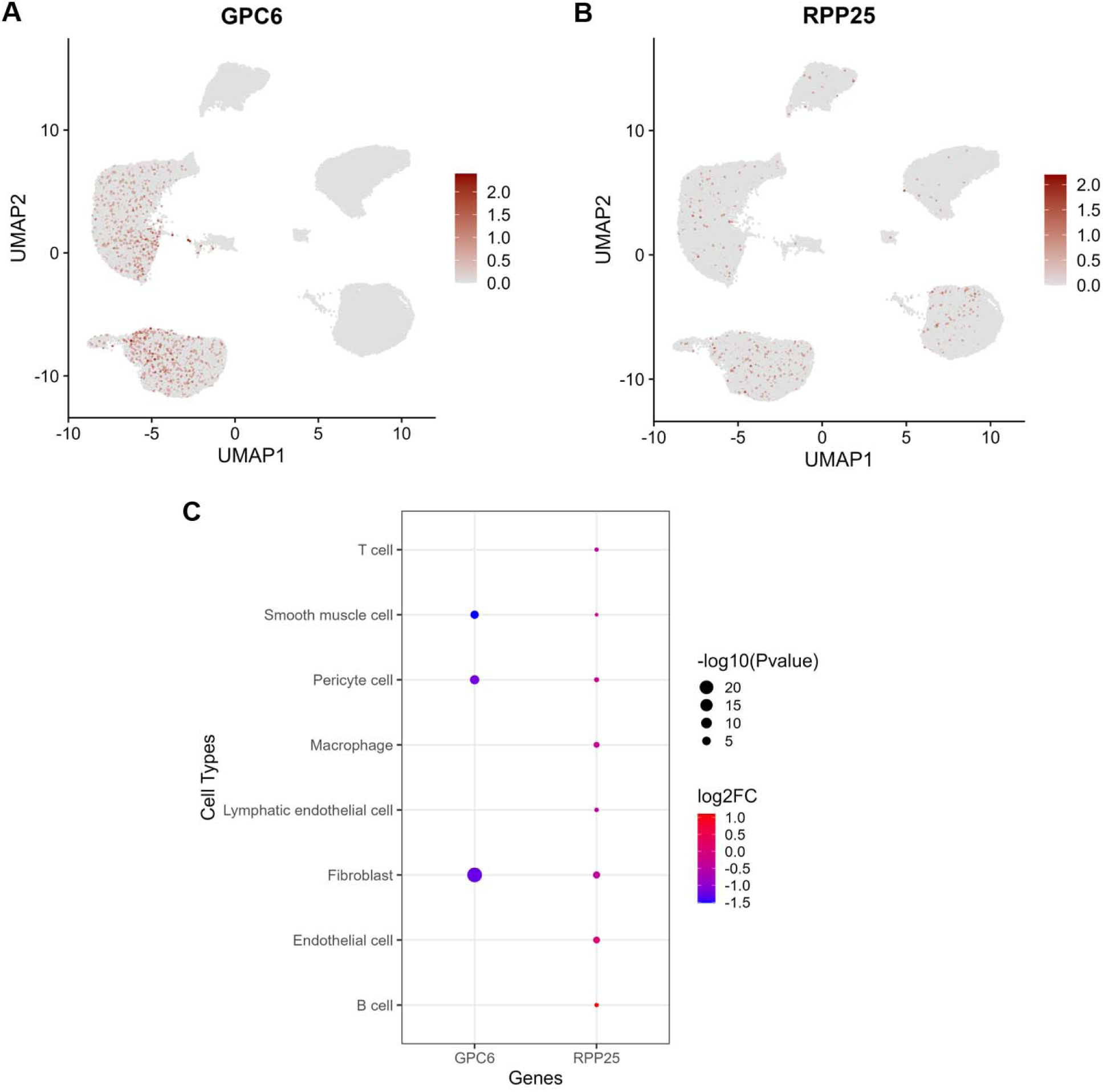
Single-cell-based differential expression analysis of GPC6 and RPP25. Feature maps show (A) GPC6 and (B) RPP25 expression profiles in human coronary artery single-cell transcriptomics. (C) Both genes show differential expression effects in atherosclerosis in specific cell types derived from human coronary artery, adding functional support to the GWAS variant-target gene relationship identified by epigenomic, chromatin, and transcription factor binding experimental data in the same tissue.

## DISCUSSION

Coronary atherosclerosis has a rich history of epidemiologic, mechanistic, histopathologic and therapeutic study, spanning population-based risk factor studies, models of atherogenesis^53^, coronary structure-functional studies^54^, imaging^55,56^, and population-level genetic risk loci and phenotype characterization^57–61^. These studies have informed the current treatment and prevention of coronary disease with cardiometabolic-inflammatory contributors, leading to metabolic (e.g., lipids, glucose, select lipoproteins) and inflammatory (e.g., statins, IL-1b antagonists) targets to intercept pathogenesis and prevent clinical events and premature cardiovascular death. Nevertheless, off-target effects (e.g., sepsis with IL-1b antagonists) and high residual clinical risk despite therapy suggest precise identification of cellular mechanisms responsible for early coronary pathogenesis may further refine novel therapeutic development. Unfortunately, translational studies in humans at a molecular level have largely been limited to quantifying circulating molecules or genetic liability for atherosclerotic CVD, missing tissue-specific contributions secondary to the inability to obtain invivo samples. Approaches to phenotyping molecular states in human coronaries are rare^52,62–64^, likely missing opportunities to prioritize important, potentially functional mechanisms of coronary atherosclerosis for intervention and tracking.

Here, we unite population-wide circulating proteomics, non-invasive coronary imaging, and coronary artery tissue-specific functional genomics and transcription to prioritize targets for CVD susceptibility. Our studies within a large population sample (CARDIA) indexed a broad human proteome to prevalent and incident CAC—an independent marker of subclinical disease and incident clinical CAD and death prior to age 60 years^65^—implicating pathogenic mechanisms across oxidative stress and lipid metabolism, extracellular matrix remodeling, immune cell function, coagulation, and inflammation^7,66,67^. Several proteins displayed evidence of causal relevance to CVD outcomes and phenotypes across multiple genetic approaches (MR and PWAS). Given our rationale that coronary artery-specific contextualization may focus discovery for CVD, we next performed the largest human coronary artery TWAS of CAC, mapping bulk transcriptomics in 268 human coronary arteries to the largest CAC GWAS, demonstrating enrichment for genes implicated by proteomic associations across similar pathways of metabolic-inflammatory activation expressed in key cell types relevant to coronary disease progression. Finally, we used coronary artery-specific regulation in MR for targets with shared evidence across proteomics and coronary TWAS, yielding a final set of 8 genes. Of note, several genes had prior biological annotation in CVD-relevant pathways, but none had been previously targeted in CVD. Phenome-wide association studies and single cell transcriptomics in human coronary arteries with and without atherosclerosis provided supportive tissue and phenotypic context. Collectively, these results not only provide the largest scientific resource to date of population-level multi-omics, coronary TWAS and coronary functional genomics-based discovery: more generally, they furnish a framework to decipher disease-relevant targets through integration of human genetic approaches with multi-omics.

Over the last decade, two parallel innovations in human genetic research have accelerated progress beyond traditional GWAS approaches: (1) broad molecular characterization of disease states (“multi-omics”) and (2) functional and tissue-based genetic approaches (TWAS, functional genomics). These approaches have leveraged the power of human genetics to identify novel disease targets through surrogate phenotype-proximal molecular traits (e.g., protein or metabolite QTLs) and to colocalize disease GWAS findings with tissue expression QTLs. Despite successes in metabolic and infectious disease, application of these approaches to human CVD has been limited. The current study comes in the context of an emergence of approaches to integrate tissue and biofluid domains to foster discovery in precision cardiovascular medicine^68^. Modern multi-omics linking genetics, transcriptomics, and proteomics have led to discovery in cardiovascular science^36,68,69^, including potential for druggability.

We predicated our approach on recent studies linking the human proteome to tissue-specific transcriptional states to implicate clinical-functional biomarkers of coronary disease^68,69^. In addition to well-known mechanisms of inflammation and fibrosis (**Table 2**), proteomics identified directionally consistent targets with strong evidence in model systems (less in humans), including *TREM2* (attenuates macrophage uptake of oxidized lipids, limiting progression of atherosclerosis in mice^21^), *Notch1* (loss potentiates *RUNX2*-mediated calcification^17^), and *ARHGAP36* (overexpression associated with connective tissue to bone transformation^18^), among others (**Table 2**). Importantly, we observed a strong concordance between effect estimates for <u>prevalent</u> and <u>10-year incident CAC</u> in CARDIA with strongest agreement for both known (e.g., MMP-12, GDF-15) and emerging mediators of vascular risk. These results were further resolved with two complementary genomic approaches, standard MR and PWAS. Standard MR approaches rely on identification of *cis*-pQTLs for causal inference, while PWAS generates optimally predictive genetic models of protein abundance not necessarily reliant on specific pQTL identification. Standard MR approaches suggested directional consistency in genomic-proteomic effects for many targets (**Figure 3A**), many of which have not been extensively implicated in human coronary disease by GWAS studies but have relevant metabolic-vascular mechanisms (e.g., *HS6ST3*^70^, *PTPRD*^71^, *CTQT1*, *DNJB9*^72^). PWAS in UK Biobank pinpointed four additional proteins with strong evidence for mechanistic involvement in atherosclerosis (*PCSK9*^31,32^, *FGFR1*, *HTRA1*, *APOC1*^33,34^) and myocardial infarction (*PCSK9*, *INHBC*^35^).

A key innovation in our approach is tissue-specific discovery via transcriptome-wide association studies in human coronary artery tissue. Our TWAS approach has recently been used to identify targets essential to cardiometabolic therapeutics^73^, though its application to coronary arteries has—to our knowledge—never previously been performed. The current report represents the largest TWAS in human coronary arteries to date. Human coronary TWAS associations with CAC included known (*PHACTR1*^37^, *MRAS*^37^*, MORF4L1-ADAMTS7*^38^) and not widely implicated targets (*GIGYF1*, *DMPK*, *RPL9*). We observed enrichment, among CAC-associated circulating proteins, for coronary artery TWAS, with targets implicated in vascular homeostasis (*NOTCH3*^44^, *TNFSF12*^45^, *S100A12*^46^) as well as several targets not previously linked to coronary disease. Through extensive MR analyses for robust causal inference, all 8 overlapping targets implicated by both TWAS and circulating proteomics had supporting causal relation to CAC or myocardial infarction. PheWAS of these genes implicated vascular, inflammatory, and obesity-related phenotypes. Importantly, none of these genes had been therapeutically targeted in CVD, with a handful implicated in CVD-adjacent mechanisms (as described above^44–46^).

Using an epigenomic atlas of regulatory elements in coronary artery and chromatin conformation capture (Hi-C) experimental data that characterize the spatial organization of chromatin, we found substantial support for the role of *trans* (distal) genetic regulation, by GWAS-implicated CAC variants, of 2 of the 8 genes. Notably, we found confirmation of the gene regulatory relationship by showing that the gene targets of the GWAS-implicated CAC variants, *GPC6* and *RPP25*, were significantly differentially expressed in single-cell transcriptomic analysis of human atherosclerosis. Neither of these have been previously implicated in CAC, though both are implicated in CVD-relevant mechanisms: *GPC6* blocks canonical Wnt signaling^48^ (critical to vascular calcification^49,50^), and *RPP25* is implicated in cardiometabolic disease^51^ and in scleroderma (a disease marked by multi-organ calcification). These results establish a comprehensive paradigm beyond traditional GWAS, leveraging the power of functional genomics, tissue-specific transcription, and population-level proteomic studies to hone disease targets.

Our study has several important limitations. While CAC is a marker of early CVD, studies across even earlier disease states reflecting the earliest changes in endothelial function (e.g., measures coronary flow and structure^55^) may uncover additional mechanisms of coronary physiology. In addition, coronary artery utilized for functional genomic approaches in our work may necessarily not reflect the range of coronary phenotypes relevant to disease discovery.

Nevertheless, concordance with signals from the circulating proteome and CAC GWAS support external validity, generalizability, and broad biological significance of our findings. Moreover, availability of coronary arteries early in the development of CVD (with appropriate preservation for molecular studies) remains challenging. We leveraged genetic instruments of the circulating proteome through PWAS, and human coronary specific PWAS is likely to yield additional mechanistic insights. Finally, discovery across heterogeneous populations may afford further generalizability. Nevertheless, multi-level genetic concordance in tissues and populations provide a focus for calcification mechanisms and biology for wider studies and therapeutic approaches.

In conclusion, we present an integrated approach to address coronary artery disease susceptibility across the circulating proteome, coronary artery-specific transcription (268 human coronaries and single cell data), and coronary artery-specific functional genomics, implicating not previously reported genes in causal CVD pathogenesis. To our knowledge, this tissue-specific genetic approach has not previously been leveraged for human genetic discovery.

Targets identified by our approach (*NOTCH3, SPINK2, S100A12, RPP25, OAF, HS6ST3, TNFSF12, GPC6*) enjoyed multi-level support, including via single cell transcription from atherosclerotic human coronary arteries, phenome-wide association, and implication in biologically relevant, CVD-adjacent mechanisms. Strikingly, the approach allowed “de-orphanization” of two loci significant in CAC GWAS without linkage to a *cis* target (*GPC6*, *RPP25*), demonstrating the power of the multi-omic approach in discovery. These results support a broader utilization of this approach as an adaptable framework applicable to all organ systems to parse precision targets for prevention, surveillance, and therapy of cardiovascular disease.

## MATERIALS AND METHODS

### Clinical cohorts

#### CARDIA

A total of 2,971 participants with circulating proteomics and CAC scores by computed tomography at the Year 25 visit were included^74^. Clinical-demographic data was collected by standardized assessment as described^65,75^, and computed tomography for CAC was performed as previously noted via standardized protocols^76,77^. In addition, 793 participants had additional computed tomographic CAC data collected at Year 35 in CARDIA for analysis of incident CAC development by Year 35 (in those individuals without CAC at Year 25). Study participants provided written informed consent, and the study was approved by the respective clinical site Institutional Review Board.

#### FHS

We studied 573 individuals with extant proteomics and computed tomographic CAC measurements from the FHS Generation 2 (“Offspring”) cohort to assess for replication of our findings in CARDIA^78^. Of note, CAC measures and proteomics were separated by median of 17.4 years in FHS. Methods for clinical and calcification traits have been reported^79,80^. All FHS participants gave written informed consent, and all study protocols received approval from the Institutional Review Board at Boston University Medical Center.

### Quantification of the circulating proteome

In CARDIA, we used the SomaScan platform (aptamer-based technology) to quantify 7,228 aptamers in CARDIA (Somalogic, Boulder, CO), as reported in previous work^74^. We excluded 71 aptamers with a coefficient of variation exceeding 20%. After log_2_ transformation and standardization of aptamer levels, aptamer values over 5 standard deviations from the mean value were winsorized. In FHS, a total of 1,128 Somascan aptamers was utilized, and methods for handling proteomic data (batch pooling, transformations, and rank normalization) have been reported^81^, and we have reproduced them with minimal change to maximize reproducibility. In brief, due to variations in collection batch in FHS, protein levels were standardized within each of the two batches, pooled, and subjected to rank normalization across all samples. Subsequent residualization against the assay plate was performed before analysis to eliminate plate-based effects^81^. Matching of CARDIA to FHS aptamers was performed by SomaScan seqID (1035 overlapping aptamers).

### Human genetic studies

**pQTL and GWAS specification:** For the target proteins, we identified protein quantitative trait loci (pQTLs, defined here as genetic instruments associated with circulating protein levels at genome-wide significance) that were measured with 4,907 aptamers in 35,559 Icelanders (ecode.com/summarydata/; quality control reported previously^82^). We conservatively selected pQTLs exceeding a genomic threshold (P<5×10^-8^) for the target proteins, leveraging both *cis*-pQTLs (+2 Mbp on both directions of the encoding locus) and *trans*-pQTL (when available). This subset was subject to linkage disequilibrium (LD) pruning (*R*^2^<0.001, 10kbp window, 1000 Genome European reference). We restricted pQTLs to those exceeding an F statistic over 10 to minimize downstream weak-instrument bias. Of 131 unique proteins that were associated with presence and extent of CAC in CARDIA, we identified at least one pQTL for 100 proteins.

For disease states, we selected three of the largest GWASs capturing CAC-relevant clinical disease states for assessment in Mendelian randomization: (1) coronary calcification (N=36,720 Europeans)^36^; (2) coronary artery disease (CAD; N=1,165,690 Europeans)^83^; (3) myocardial infarction (N=639,221 Europeans)^84^. A summary of studies is presented **in** Supplementary Table 4.

#### Two-sample Mendelian randomization (MR)

Our two-sample MR framework tested whether proteins implicated in our initial epidemiologic and tissue evaluation were causal by human genetic approaches to CAC and relevant outcomes. We used pQTLs (as instrumental variables [IVs] for genetically determined protein levels) as exposures and our three CAC-relevant clinical disease states as outcomes. Quality control was performed using TwoSampleMR R package^2^. We primarily tested potential causal associations with inverse variance weighted robust-penalized method, using MendelianRandomization R package^3^. We used ‘default effect’ model for association testing, or ‘random effect’ model when IVs for the protein demonstrated significant heterogeneity in effect size (Q-het P<0.05). Given the multiple lines of evidence preceding these approaches, we considered the protein as potentially causally associated with the CAC-relevant outcome at a nominal P <0.05. In addition, we used median penalized (to limit IV-related bias) and Egger-penalized robust methods (for pleiotropy) as sensitivity analyses to enhance our confidence in causal interpretations, consistent with STROBE-MR guideline for causal inferencing using observational data^85^.

### Single-cell RNA sequencing of human coronary arteries

We leveraged single-cell RNA sequencing (scRNA-seq) data derived from 13 human coronary arteries (8 with atherosclerotic lesions and the remainder lesion-free controls) involving 56,183 cells^52^. We analyzed the data using the Seurat R package^86^. Doublet removal was performed using scDblFinder^87^. As ambient RNA contamination can negatively affect gene expression profiling, we performed correction via DecontX using default parameters^88^. We applied raw count normalization through a regularized negative binomial regression approach as implemented in SCTransform^89^. To avoid confounding by cell cycle state, we adjusted for cell cycle variance. For integration of processed sequencing libraries across potential technical differences and dataset-specific conditions, we used Harmony^90^. We applied dimensionality reduction using principal component analysis (PCA) on the normalized counts, leveraging the first 30 principal components (PCs) for clustering. We invoked the functions RunUMAP (using 30 neighboring points for local approximation of manifold structure) and FindNeighbors (using k= 20 for the *k*-nearest neighbor algorithm) in Seurat, setting reduction to ‘harmony’ to use the Harmony embeddings. Cell type annotation leveraged transfer learning from the Tabula Sapiens (specifically vasculature data in 42,650 cells available at https://cellxgene.cziscience.com/e/a2d4d33e-4c62-4361-b80a-9be53d2e50e8.cxg/)^91^. For differential expression analysis between the samples with atherosclerotic lesions and the lesion-free samples, we used pseudobulking-based DESeq2^92^.

### Proteome-wide association studies

We performed proteome-wide association studies (PWAS) of CAC, using genetic models of circulating proteome developed in a large-scale proteo-genomic study (N = 7,213, European ancestry)^93^. The models had been trained using the PrediXcan methodology applied to 4,657 aptamers (Somalogic) representing 4,435 unique genes. We included only those proteins that attain nominal significance during model training (P<0.05, cross-validation *r*=0.10, 1,340 unique proteins). This resource does not overlap with the UK Biobank (N = 361,194), allowing us to estimate the association between the genetically determined circulating protein expression and related traits in an independent phenomic resource. We used GWAS summary statistics from a linear model of a phenotype as a function of the first 20 genotype-based PCs, sex, age, age^2^, sex*age, sex*age^2^, and the protein under test to estimate the effect size of the protein on the two phenotypes in the UK Biobank: coronary atherosclerosis (code I9_CORATHER) and myocardial infarction (code I9_MI). Of 753 proteins associated with presence and extent of CAC in CARDIA (derivation P<0.05), 316 were tested in PWAS.

### Transcriptome-wide association studies

We developed gene expression models in human coronary artery samples from 268 unrelated individuals. We used our JTI TWAS methodology, which resulted in a substantial increase in the number of imputable genes (niGenes = 9,918) relative to the PrediXcan methodology (niGenes = 5,050). We trained prediction models of normalized gene expression using sex, platform, and 5 genotype-based principal components as covariates. The features consisted of common SNPs (minor allele frequency > 5%) in the gene’s *cis*-region whose extent, a model hyperparameter, was determined using cross validation. The tissue-specific JTI association test controlled the type I error rate: at the significance level of 5%, the type I error rate was 4.96% (compared to 5.04% for PrediXcan and 5.17% for UTMOST). We also performed phenome-wide association studies (PheWAS) for a set of prioritized genes by identifying the associations of the genes with the heritable phenome (heritability h^2^ P < 0.05) as captured in the UK Biobank.

### Causal inference on TWAS associations

To provide additional support for the role of the implicated genes in CAC, we performed causal inference via MR with gene expression as “exposure” and CAC as “outcome”. For inference on causality, we used five MR approaches, each relying on strong assumptions on the underlying ground truth: MR-Egger^94^, median-based^95^, maximum likelihood^96^, inverse-variance weighted^97^, and MR-JTI^98^.

### Functional genomics

We utilized a broad collection of 18 assays for the identification of human coronary artery regulatory elements. These assays measure chromatin marks: H3K27ac, H3K27me3, H3K4me1, H3K4me2, H3K4me3, H3K36me3, H3K79me2, H3K9ac, H3K9me3, and H4K20me1; chromatin accessibility, specifically DNase-Seq and ATAC-Seq; and transcription factor binding (including cohesin complex subunits RAD21 and SMC3 and RNA polymerase II subunit POL2RA). Leveraging these epigenomic, chromatin accessibility, and transcription factor assays within a regulatory annotation framework^99^ allowed us to classify the regulatory elements (e.g., active enhancers, distal or proximal insulators, active promoters) in human coronary artery. We leveraged tissue-specific Hi-C data to identify the target gene of a regulatory element in coronary artery.

### Associations of the circulating proteome to CAC

In CARDIA, protein aptamers were log transformed, centered, standardized to unit variance, and winsorized to 5 standard deviations for regression. Given absence of known datasets with harmonized proteomic platform and concurrent measures of CAC, we randomly split our CARDIA sample into a derivation (70%) and validation subsample (30%). We estimated models in the discovery subsample for presence/absence of CAC (as a binary variable in logistic models) and its extent (continuous in linear models; modeled as ln[CAC+1] to account for zero CAC values). While ln(CAC+1) transformation does not fully normalize the distribution, it is a commonly used approach in prior studies to accommodate the skewed nature of CAC data for statistical modeling^36,57,100^. Each model contained one aptamer at a time, adjusted for age, sex, and race, with a 5% false discovery rate (Benjamini-Hochberg FDR) applied across models to control for multiplicity. Aptamers that were significant at a 5% FDR threshold were passed to identical models in our validation subsample. Aptamers that were significant at 5% FDR for both presence of CAC and its extent in both derivation and validation subsamples, were analyzed in similar logistic regression models (adjusted for age, sex, and race) for incident CAC among subjects who had CAC scores of 0 at Year 25 and available computed tomography data at Year 35. Aptamers in the incident CAC models were considered significant using a threshold of 5% FDR.

In order to explore generalizability of our findings across geographies and populations, we studied population-level proteomics in FHS. In FHS, protein profiling was completed in two batches. In batch one, 1129 aptamers were profiled in 821 individuals, and batch 2 included an expanded panel of 1372 aptamers, which was assayed in 1092 participants. Aptamer levels of 1128 aptamers common to both batches were log-transformed, standardized within batch, then pooled and normalized to a mean value of 0 and SD unit of 1 using the Blom rank-based inverse-normal method. These values were then regressed on assay plate ID to account for plate effects, and the standardized residuals were used for regression. Replication studies in FHS were performed in a similar fashion (logistic for presence or absence of CAC; continuous linear for extent as standardized ln[CAC+1]), with each protein in separate models adjusted for age, sex, and race (white vs. non-white) (with 5% FDR used to control multiplicity). We assessed concordance of CARDIA and FHS effect size via Pearson correlation.

## Data Availability

No new code was developed as part of this study. Data from CARDIA used in these analyses are available through the Coronary Artery Risk Development in Young Adults study (CARDIA; cardia.dopm.uab.edu) or at dbGaP (for proteomics, dbGaP identifier phs003491.v1.p1). Data from the Framingham Heart Study (FHS) is available at dbGaP (dbGaP identifier pht006013) or via contact with the FHS coordinating center (www.framinghamheartstudy.org). UK Biobank data (accessed under application number 94960) is available at UK Bio bank Research Access Portal. Code used for analysis in this study are available on Zenodo (https://doi.org/10.5281/zenodo.3842289) and on the Github repository for this project (https://github.com/gamazonlab/CACOmics).

## Acknowledgments

CARDIA is conducted and supported by the National Heart, Lung, and Blood Institute (NHLBI) in collaboration with the University of Alabama at Birmingham (75N92023D00002 and 75N92023D00005), Northwestern University (75N92023D00004), University of Minnesota (75N92023D00006),Kaiser Foundation Research Institute (75N92023D00003) and by grant R01-HL098445 (NHLBI) to Vanderbilt University and Wake Forest University. Dr. El-Sabawi is supported by the National Institutes of Health (NIH) (T32 HG008341). Dr. Amancherla is supported by an American Heart Association (AHA) Career Development Award (#929347), the NIH (K23HL166960), the Red Gates Foundation, and an International Society for Heart and Lung Transplantation Enduring Hearts Transplant Longevity Award. Dr. Gamazon is supported by R01HG011138 (NHGRI) and U01DK140952 (NIDDK). Dr. Shah is supported by grants from the NIH.

## Author contributions

BE, MB, PL, XH, NK, MY, PG, BLAP, ASP, QS, SZ, LS, EFE, JL, PE performed analysis. LC, GL, JT, JC, DLJ, RK collected data (proteomics or clinical). EG, KN, JB, SK, QS, MN, RS supervised analysis. BE, MB, PL, EG, RS wrote the manuscript. All authors interpreted data and edited the manuscript.

## Supplementary Figures

**Supplemental Figure 1.**
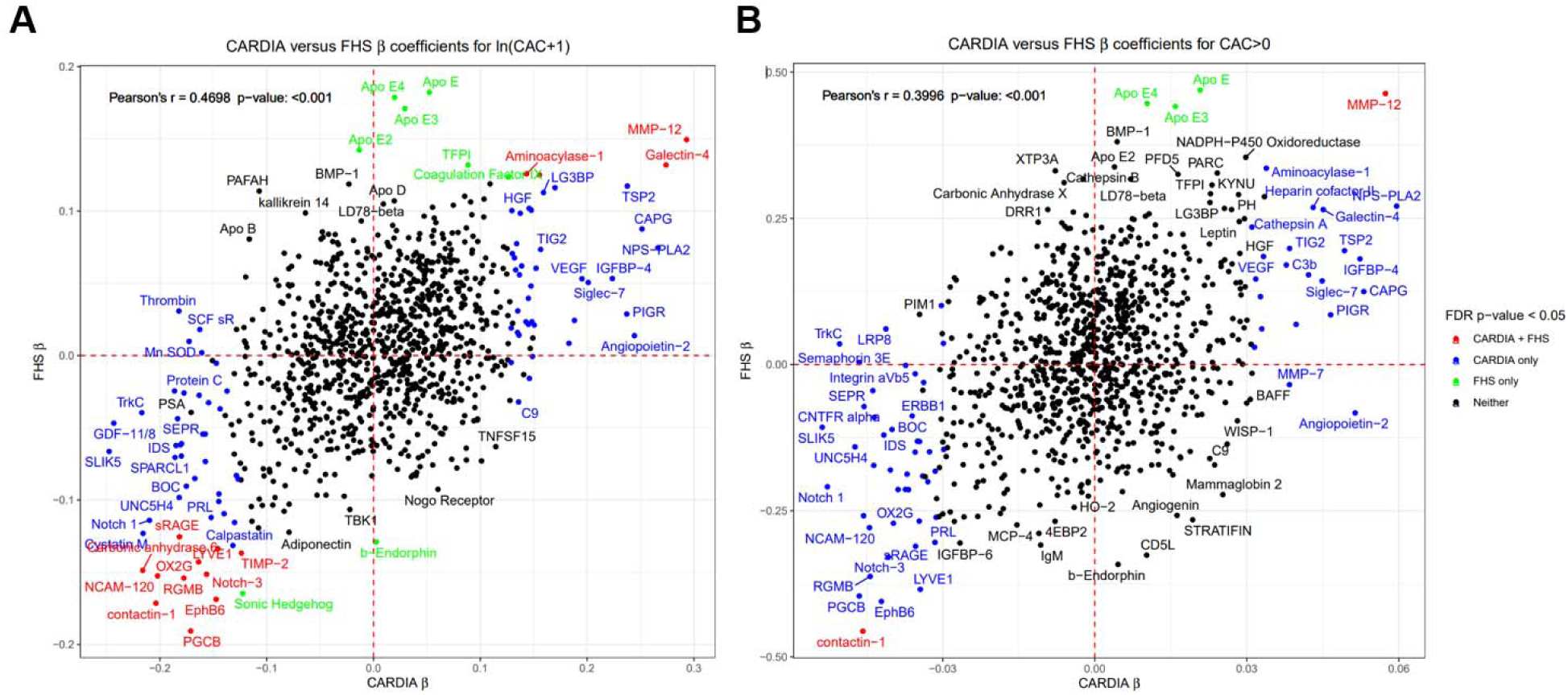
Replication of proteomic associations with CAC in the Framingham Heart Study. (A) Scatterplot showing aptamer effect sizes from the derivation sample in CARDIA versus the Framingham Heart Study (FHS) for ln(CAC+1). (B) Scatterplot showing aptamer effect sizes from the derivation sample in CARDIA versus FHS for CAC>0.

## Supplemental Tables Legends

**Supplemental Table 1.** Full list of regressions of individual aptamers with ln(CAC+1) and CAC>0 in CARDIA at Year 25.

**Supplemental Table 2.** Regressions of 136 individual aptamers that were related to prevalent CAC>0 and extent of CAC at Y25 with incident CAC in CARDIA at Year 35.

**Supplemental Table 3.** Regressions of individual aptamers with ln(CAC+1) and CAC>0 in the Framingham Heart Study.

**Supplemental Table 4.** Characteristics of genome wide associations studies.

**Supplemental Table 5.** Full results of 2-sample Mendelian randomization.

**Supplemental Table 6.** Full results of protein wide association studies (PWAS) for coronary artery disease or myocardial infarction.

**Supplemental Table 7.** Full results of coronary artery-specific transcriptome-wide association studies (TWAS) for CAC.

